# Health care provider and client experiences of counselling on depot medroxyprogesterone acetate subcutaneous (DMPA-SC) for self-injection in Malawi

**DOI:** 10.1101/2023.05.24.23290478

**Authors:** Chelsey Porter Erlank, Gracious Ali, Frehiwot Birhanu, Melinda Stanley, Jessie Salamba Chirwa, Fannie Kachale, Andrews Gunda

## Abstract

Since the introduction of depot medroxyprogesterone acetate (DMPA-SC) in 2018, Malawi has achieved national coverage of trained providers in the public sector and steady increases in uptake of DMPA-SC. However, the rate of clients opting to self-inject DMPA-SC has remained lower than early acceptability studies suggested. Providers play an instrumental role in building client confidence to self-inject through counselling. This cross-sectional qualitative study explored the perspectives of providers and injectable clients on the integration of self-injection into contraceptive counselling, to identify best practices and gaps. The study was conducted at public sector sites in three districts (Nkhotakota, Mzimba South, Zomba) in Malawi. In-depth interviews were conducted with provider-administered injectable clients, self-injecting clients, and DMPA-SC trained providers. All providers interviewed reported successfully integrating self-injection into their approach. During health education sessions, providers tended to focus mainly on benefits of self-injection to spark interest in the method, and then follow that up with more in-depth information in individual counselling. Due to time pressures, a minority of providers reported replacing individual counselling with small-group counselling and limited their use of peer testimonials, visualizations, and demonstrations. Most providers skipped client practice on inanimate objects, feeling this was either not necessary or not appropriate given stock or resource constraints. Current self-injecting clients showed the best recall for self-injection steps and tended to report having received comprehensive, supportive counselling including aspects such as peer testimonials, visualizations, and demonstrations to build confidence. Injectable clients who had declined self-injection tended to demonstrate less detailed recall of key self-injection messages and report receiving incomplete information, and lack of peer testimonials, visualization, or demonstrations. Comprehensive counselling and training from supportive providers, including best practices identified in this study, are vital to improving client confidence to self-inject. Providers should be supported to overcome time- and resource-pressures to invest in these best practices.

## Introduction

Injectable contraceptives are the most popular method used in Malawi – making up almost half (49.8%) of the contraceptive method mix. [1] A new subcutaneous formulation (DMPA-SC) of the popular injectable depot medroxyprogesterone acetate intramuscular (DMPA-IM) was introduced in Malawi in 2018. DMPA-SC is safe, highly effective at preventing pregnancy, and administered every three months. DMPA-SC differs from DMPA-IM in that it comes in a pre-filled, ‘all-in-one’ Uniject syringe, it is injected subcutaneously, and contains a lower dose of DMPA. The Uniject syringe means DMPA-SC can be administered by any trained person, including community health workers, pharmacists and contraceptive clients themselves, where it is registered for use by these groups.

Research on DMPA-SC in Malawi has shown high acceptability rates for DMPA-SC being administered ‘at home’ as opposed to in a clinic by a provider (70% among injectable users); high rates of willingness to continue to self-inject (SI) among trained clients (98%); and high rates of willingness to SI in the future among provider-administered (PA) injectable clients (78%). [2] The SI option has also been found to be associated with increased continuation of contraceptive use among injectable clients (73% SI clients continuing at 12 months compared to 45% PA clients). [3] Qualitative studies in Malawi have demonstrated preference for DMPA-SC over DMPA-IM among both providers and clients, due to the ease of administration and the time- and travel-savings associated with SI versus PA. [4] Based on these positive results, the Ministry of Health (MOH) approved the national roll-out of DMPA-SC (both PA and SI) in 2018. By 2021, all public sector facilities nationally had at least one DMPA-SC trained provider.

Since the national scale up, analysis of Health Management Information System (HMIS) data shows that uptake of DMPA-SC has steadily increased. However, the rate of new injectable users choosing the SI option has remained lower than rates indicated by early acceptability studies. [2, 5] For example, in Q4 2020 only 12% of DMPA new injectable users took up the SI option (versus 38% and 50% taking up DMPA-SC PA and DMPA-IM respectively). SI accounted for just a quarter (25%) of DMPA-SC uptake in Q4 2020, and continued to average around 21% in subsequent quarters. [5]

During supervision visits to facilities across Malawi in 2020-2021, the MOH observed occasional provider bias against SI, with some providers reporting it took too long to train women to SI. Qualitative evidence from Malawi shows training women on SI may take longer than PA options, due to the addition of the SI demonstration, client training and practice sessions on top of the standard method-specific counselling on advantages and side effects. According to SI clients in one study in Malawi, SI counselling and training took approximately 20 minutes, while providers reported 27 minutes on average (10-60 minutes range). [4] Exactly *how* the SI option is introduced by providers can also influence client confidence to take up this option – for example, a recent study found that standardizing messaging on SI (including specific reassurances about the common SI concerns) was associated with higher SI uptake. [6] A recent study in Uganda also found that training quality may affect adolescents’ confidence to SI independently, [7] while mixed-methods research conducted by the Delivering Innovation through Self-Care (DISC) program in Nigeria and Uganda has identified that providers can ‘gatekeep’ the SI option (i.e. introduce barriers to information and support to SI for some women) based on their own biases or lack of confidence with the product. [8] This study was designed to investigate barriers and enablers of self-injection uptake more broadly, with a specific objective to investigate best practices and potential gaps in providers’ approaches to integrating the SI option into their contraceptive counselling for new and returning injectable users in Malawi.

## Materials and methods

MOH Malawi and Clinton Health Access Initiative (CHAI) collaborated in 2021 to conduct a cross-sectional qualitative study investigating the barriers and enablers to DMPA-SC SI uptake in Malawi, including a specific emphasis on understanding the contraceptive counselling interaction between providers and clients, from both perspectives. Specific outcomes of interest included: provider perspectives on enablers and barriers of integrating DMPA-SC intro their contraceptive counselling approach; client perspectives on the information received about DMPA-SC from providers; client perspectives on the quality of training and support provided during self-injection training; provider and client perspectives on the length of time counselling on DMPA-SC takes. The results of a secondary research question on perspectives of adolescents with unmet need for contraception on DMPA-SC SI are published elsewhere. [9]

Study sites were six randomly-sampled public sector facilities in three districts (Nkhotakota, Mzimba South, Zomba) – one district from each of Malawi’s three regions. Districts and facilities were sampled randomly to minimize selection bias, however only public facilities that provided 10 or more DMPA-SC services per month were included in the sampling frame, to ensure enough injectable clients could be sampled. At each site, providers and injectable clients were purposively sampled according to inclusion criteria (for providers, this meant having been trained in DMPA-SC; for clients, this meant having used either a self-injected or provider administered injectable contraceptive in the last nine months).

Data was collected using semi-structured in-depth interview guides. Several participatory activities were integrated to elicit detail about the provider-client counselling interaction. Firstly, providers were asked to role-play contraceptive counselling and SI training with the data collectors acting as ‘clients’. Providers were reassured that this exercise was purely to understand variations in approaches between providers, and that their responses would be anonymized, to try and minimize possible Hawthorne effect. Secondly, recognizing that some clients may not always critically appraise the quality of their care when they are unaware of the standards expected, [10] injectable clients were shown one of two videos to establish a standard against which they could assess the quality of the counselling and SI training received:

1. Video 1 showed two actors demonstrating a best practice counselling and SI training session between provider and client (S1 contains the script for the video). This video was created for the study and approved by the MOH for use for study purposes. It was shown to PA injectable clients who had chosen not to take up the SI option to help them critically compare to their own experience of counselling.
2. Video 2 was developed by PATH International [11] and explains the critical SI information that a self-injecting client needs to know. This animated video was shown to current/recent SI clients to help them identify any gaps in their knowledge.

As Nkhotakota and Zomba are predominantly Chichewa-speaking districts, while Mzimba South is predominantly Tumbuka-speaking, all study tools and videos were translated or dubbed into both languages for study purposes.

The final sample size was based on theoretical saturation, which refers to the point at which no new ideas emerge from a sample diverse in relevant characteristics and experiences. [12] Clear variation in themes emerging from the two DMPA client populations (PA and SI) during early data collection led to the decision to slightly expand sample sizes in those two groups to ensure saturation could be reached. The final sample of 24 providers included 18 HSAs and 6 facility-based providers, most of whom received their DMPA-SC training a year or more ago. The final sample of 40 clients included 15 clients who had self-injected in the last nine months and 25 clients who had received a provider-administered injectable (DMPA-IM or DMPA-SC) in the last nine months. Most clients were aged between 20 and 39 years, were married and had 1-2 children. Full details of the provider and client sample characteristics are outlined in Table 1.

**Table 1.** Participant characteristics

| Provider sample (N=24) | Facility-based providers | Health Surveillance Assistants (HSAs) |
| --- | --- | --- |
| **Time since trained** |  |  |
| Less than a year ago | 2 (33%) | 4 (22%) |
| A year or more ago | 3 (50%) | 11 (61%) |
| Unknown | 1 (17%) | 3 (17%) |
| **Total** | **6 (100%)** | **18 (100%)** |
| **Client sample (N=40)** | **DMPA-IM / DMPA-SC provider-administered clients** | **DMPA-SC self-injection clients** |
| **Age** |  |  |
| 18-19 years old | 6 (24%) | 4 (27%) |
| 20-29 years old | 9 (36%) | 5 (33%) |
| 30-39 years old | 8 (32%) | 3 (20%) |
| 40-49 years old | 1 (4%) | 2 (13%) |
| Unknown | 1 (4%) | 1 (7%) |
| **Marital status** |  |  |
| Never married | 3 (12%) | 1 (7%) |
| Married | 18 (72%) | 12 (80%) |
| Divorced/separated/widowed | 3 (12%) | 1 (7%) |
| Unknown | 1 (4%) | 1 (7%) |
| **Children** |  |  |
| No children | 1 (4%) | 0 (0%) |
| 1-2 children | 16 (64%) | 9 (60%) |
| 3-4 children | 6 (24%) | 5 (33%) |
| Unknown | 2 (8%) | 1 (7%) |
| **Method used in last 9 months** |  |  |
| DMPA-IM | 20 (80%) | N/A |
| DMPA-SC provider-administered | 5 (20%) | N/A |
| DMPA-SC self-injected | N/A | 15 (100%) |

| Heard of SI before the in-depth interview |  |  |
| --- | --- | --- |
| Yes | 20 (80%) | 15 (100%) |
| No | 5 (20%) | 0 (0%) |
| Total | 25 (100%) | 15 (100%) |

All data was collected in October 2021. No identifying information about participants was collected other than broad descriptors relating to basic demographics or cadre. Quantitative descriptors were collected on paper by the study team and manually entered into Excel for descriptive analysis (Table 1). Qualitative data was collected by trained research assistants, audio-recorded, transcribed verbatim and translated into English for analysis. Data was coded deductively by three qualitative researchers using Dedoose software, then code reports (S2) exported from Dedoose for thematic analysis. For some themes (for example, the counselling messages covered during provider role-plays), a ‘framework approach’ was used, charting the coded data into a matrix to allow comparability across in-depth interviews and against national in-service training content (provided by MOH). Where relevant, sub-group analyses were conducted by splitting code reports up between provider-administered and self-injection client groups or between HSAs and facility-based provider groups, to note any variation in themes or opinions between the sub-groups.

All participants were asked to provide voluntary written informed consent to participate in this study prior to interview. Ethical approval to conduct this research was granted by the National Health Science Research Committee (NHSRC) – an independent international review board in Malawi with Federal-Wide Assurance (IRB00003905, FWA00005976).

## Results

### Integrating self-injection into health education

All providers claimed to have integrated DMPA-SC and the SI option into their health education run-through of all the contraceptive methods (which they conducted as either a group session during a structured clinic or, for some HSAs, one-to-one with individual clients during home-based visits). Most providers said they still mentioned DMPA-SC and the SI option at this stage, even if they did not currently have DMPA-SC in stock (Malawi experienced national shortages of DMPA-SC in 2020-2021 due to global disruption of supply).

Some providers mentioned incorporating testimony from satisfied SI clients at this stage, to demonstrate to other women that it was possible for women like them to SI. Many providers reported emphasizing the benefits of SI at this stage; particularly that SI could reduce visits to facilities, which they felt would be the most appealing feature to women. One provider explained that this was to pique client interest in DMPA-SC, knowing that the details of the method would be covered later in individual counselling:

> “[In health education sessions]… we cannot focus on the bad side because she will not choose the method, but we should explain to her the advantages of the method. Then she understood [sic] and makes her choice. And then we train her on this method [later].” – HSA, Nkhotakota

Echoing this, most PA injectable clients mentioned having first heard some high-level messages on SI during either a health education session and/or sometimes through other women. For these clients, their level of recall was also typically limited to only the benefits of SI:

> “Yes, they [providers] told us that the injection is good, the self-injection. [Interviewer: Alright, did they say anything about the side effects of this injection? …] No, they did not say anything on that, they just explained about the benefits…” – married PA client, 30-39 years, Zomba

A small group of PA injectable clients reported little or no awareness of even the high-level messages about SI. These were mainly long-term DMPA-IM users who reported that they tended to skip or not pay attention during health education sessions at their re-injection visits, citing satisfaction with DMPA-IM and lack of motivation to switch methods:

> “I can’t explain [about DMPA-SC] because I did not pay attention [during health education] … [Interviewer: Why didn’t you pay attention?] It’s because I don’t want to be convinced with a new method… I did not pay attention [be]cause the one [contraceptive method] I am using works well on me, so I don’t want to hear other methods.” – married PA client, 30-39 years, Mzimba South

### Integrating self-injection into individual method-specific counselling

If clients expressed interest in DMPA-SC after health education, providers then reported offering them in-depth individual counselling where they asked questions about contraceptive history, side effects and checked for contraindications. Most providers stressed the importance of having the one-to-one space with clients to tailor counselling to their needs. However, a handful of providers reported grouping women who expressed interest in a particular method to avoid repeating themselves and to save time – this practice included but did not seem to be exclusive to counselling on DMPA-SC. During stockouts of DMPA-SC, providers ranged in their approach at this stage: from encouraging women to temporarily use other methods, to still proceeding with DMPA-SC counselling/training but limiting clients’ opportunities to practice and/or their number of take-home doses, depending on stock availability.

During counselling role-plays, most providers demonstrated comprehensive counselling on DMPA-SC messages that aligned with national in-service training content, including proactively advising on safe storage and disposal of the Unijects. Most providers were able to explain the benefits (pregnancy prevention, reduced visits to facilities, discretion, etc.) and potential side effects (e.g. menstrual changes, weight change, headaches, etc.) of DMPA-SC, relative to other methods.

However, some key messages from the national in-service training content that providers often missed during role-plays included advice on what women should do:

- if they missed their re-injection window
- if they experienced any irritation or dimpling at the injection site
- to return unused units if they decided not to use them

In role-plays with the data collectors, providers shared their techniques for familiarizing the concept of SI, for example making comparisons between DMPA-SC and other needle-based concepts women were familiar with – usually DMPA-IM, but also sometimes things like vaccines, malaria test kits, or insulin injections:

> “…we also tell them that the needle is small comparing with Depo Provera [DMPA-IM]. When you show the… the two needles, they say this is long and this small and easy just like malaria test kit. When we counsel her, they understand although they might still be afraid.” – HSA, Nkhotakota

During role plays, most providers showed good recall of the critical steps of SI from national in-service training content. The critical steps most commonly mentioned by providers during the role plays included:

- the importance of shaking and activating the Uniject
- pinching the skin at the injection site
- injecting at a downward angle
- not rubbing the injection site after injecting
- disposing of the product safely in a closed container and returning it to a heath official

A few of the self-injection steps that were less commonly recalled by providers included:

- Handwashing before self-injecting
- Checking expiration dates before self-injecting
- Removing the needle cap and not replacing it
- Pressing the reservoir for 5-7 seconds
- The specific sequencing of removing the needle and releasing the ‘pinched’ skin
- Calculating and noting down the date for the next injection

From the client side, there were striking differences between the level of recall of critical SI steps between PA and SI clients. Most PA clients recalled only high-level information (typically only shaking and activating the Uniject and pinching the skin before injecting on the thigh or stomach). By contrast, current/recent SI clients showed the most detailed spontaneous recall for most of the critical SI steps, typically able to walk through most or all the steps from memory:

> “This is how we hold Sayana [showing the interviewer using hands] then we shake it, after shaking it we press its neck and then the needle is pushed inside then we hold the place where we want to inject ourselves and then we inject the needle. The moment we realize that the needle is inside the skin then we press the medicine until all is finished then we start pulling out the needle little by little.” – divorced/widowed SI client, 20-29 years, Nkhotakota

Some PA injectable clients were positive about the quality of counselling they received, even after comparing it to the best practice counselling/training session video (S1). However, several others felt their own experience of counselling/training was less detailed than the session in the video:

> “[Interviewer: Ah why have you not tried it [self-injection]?] because I was not trained properly, I did not receive the proper training … but they also did not explain that this is how you perform self-injection very well …they also did not talk about the consequences [side effects]” – PA client, missing age and marital status, Zomba

For example, some PA clients picked up on topics covered in the best practice video that had been missing from their counselling, namely exactly how and where to self-inject, or noted that they had not received a visual aid/calendar that might have helped them remember the steps:

> “The provider said we have three places where we can self-inject but, in the video, they have said we have only two places where we can self-inject.” – married PA client, 30-39 years, Mzimba South

> “…we were not given the calendar… it was a verbal calendar explaining that after 90 days you should do it…. But here [in the video] there is a calendar [with the job aid showing the SI steps] from washing hands to self-injecting.” – married PA client, 40-49 years, Zomba,

By comparison, SI clients were more likely to say they had received complete information in their counselling, and therefore knew most or all the key information covered in the PATH video: [11]

> “… everything that we were trained [on] is in the video… After I watched the video and the training that I received from the providers… I can see that [I] am able to do everything in order and there is no problem and the providers trained us well” – married SI client, 30-39 years, Zomba

Only a few SI clients reflected that the topic of possible side effects of DMPA-SC had been covered in more depth in the video than in their original counselling:

> “The training I received from my provider is different [from the video] because [in the video] they have said that some get fat, some menstruate, some [have] stomach ache and some headache. They did not explain this.” – married SI client, 20-29 years, Nkhotakota

All SI clients reported feeling reassured enough in their counselling to proceed to being trained in SI. By contrast, only a few PA clients reported that they had decided to proceed with training in SI after receiving counselling – the majority reported that they still had too many concerns about pain or doubt in their own ability to SI to even with training.

### Conducting self-injection training

After counselling on DMPA-SC, answering client questions, and establishing no contraindications, providers reported offering women the opportunity to be trained on SI. Several providers commented that, if they had spent enough time addressing concerns during counselling, most clients would choose to be trained in SI, while only a few women would request PA instead:

> “…if you do the counselling very well, they do not have any concerns as long as they have understood every procedure on how they can self-inject and also know the next date to self-inject in doing so they will not forget.” – Facility-based provider, Mzimba South

To kick off SI training, most providers reported first conducting some sort of visual demonstration of the Uniject and SI steps. This typically involved using a visual aid (e.g. posters, job aids or the visuals on the back of SI calendars) and/or gestures and demonstrations with a real Uniject – where DMPA-SC stock availability allowed. Many providers emphasized the importance of women being able to visualize the SI steps to reassure them of the simplicity of it:

> “…most people easily understand when they are able to see the things that they are being trained in, how it is operating…” – HSA, Zomba

Providers were generally very positive about the DMPA-SC SI calendars used in Malawi (which include a visual reminder of the SI steps on the back), which they felt served multiple purposes – acting as a visual aid during training, serving as a reminder of the SI steps for women at home, and helping women keep track of their re-injection dates:

> “Like calendars they are so helpful… It contains details on the steps on how self-injection should be done and also it has some pictures that help the clients to see the process. We also use the pictures when counselling clients … It helps especially when they are alone, they are able to refer to the instruction sheet.” – HSA, Nkhotakota

After the visual demonstration, most providers then encouraged clients to practice with a Uniject. While national guidelines suggest women should practice first on inanimate objects, such as a condom filled with sugar, only a few providers talked about doing this in practice (and, if they did, they typically used cheaper materials than sugar, such as condoms filled with sand, or oranges or tomatoes). Most providers in this study instead reported encouraging women to ‘practice’ SI directly for the first time on their thighs or stomachs, under supervision. This deviation from recommended protocol was said to be due to concerns about ‘wasting’ scarce DMPA-SC stock; provider time constraints; concerns about the cost of sugar; or because providers felt women were not ‘convinced’ by practicing on inanimate materials:

> “…most of the times the women are reluctant that this [condom filled with sugar] is not a real thing, and they want a demonstration on the actual body… so we don’t rely on it too much…” – Facility-based provider, Zomba

Providers often noted that practice was the most challenging part of SI training, requiring repeated feedback to ensure women did it correctly. Some providers seemed to treat this step as something of an ‘exam’ for women, using the terms ‘pass’ and ‘fail’ to denote if they were happy or unhappy with the woman’s attempt:

> “…we compare … how they are injecting themselves and see where they are doing wrong, and we help them to improve on that. And the ones who have passed [successfully self-injected], we congratulate them and tell them to continue at home. But for the ones who have failed, we tell them.” – HSA, Nkhotakota

A few providers talked about revisiting demonstrations and training on SI over time (e.g. over the course of several PA re-injection visits) to build the confidence of women who had previously ‘failed’ practice sessions.

From the client perspective, SI clients were slightly more likely than their PA counterparts to mention having seen a visual demonstration of SI steps during SI training, while some said they had seen another woman self-inject before being encouraged to ‘practice’ on themselves. By contrast to SI clients, very few PA clients mentioned seeing a visual demonstration of any kind, nor seeing other women self-inject first before being asked to ‘practice’ on themselves. Very few clients in either the SI or PA group mentioned having practiced on inanimate objects – most said that their first ‘practice’ had been on their own thighs under provider observation:

> “We were taught how to inject ourselves; we were asked if [we] would manage and I agreed and attempted to inject myself while the HSA was watching, she confirmed that I had done it well and could manage to do it on my own.” – married SI client, 40-49 years, Zomba

> “[Interviewer: Did they give you a chance to practice] Yes, they did [Interviewer: Did you self-inject? What did you use to practice?] They gave us a chance like me I practiced on my body” – married PA client, under 20, Mzimba South

In general, SI clients reflected positively on their counselling/training and credited this with helping them overcome initial fears:

> “I had fears at first. But the fears went off because we were self-injecting under direct observation by the provider, they were instructing us on how to do it [Interviewer: What did you [do to] deal with the fears apart from providers being there for you?] It’s because of the counselling they gave us. When we started the procedure, everything was going as they had taught us, so this relieved us our fears.” – married SI client, 20-29 years, Mzimba South

Some SI clients described how providers helped them overcome their fears and build their confidence over repeated visits, if they had initially been too afraid to try SI:

> “Like for me I asked them to inject me because I was not confident. But I was told it [DMPA-SC] was meant for self-injection. During the second visit, they counselled us again and this time I understood and managed to self-inject” – married SI client, 30-39 years, Mzimba South

When asked why they had not taken up SI after training, the few PA clients who had received some SI training tended to blame themselves rather than the quality of their counselling/training. For example, they were more likely to talk about struggling to overcome initial fears; lacking confidence in their own ability; or mirroring the providers’ language about having ‘failed’ their practice:

> “I wanted to find out if I will be able to self-inject. If I will do it, I will be using that one. But I failed. [Interviewer: You failed?… How exactly did you fail?] I failed to open [activate]. They say that for it to be opened [activated], it has to make a sound and I failed to do that. I was like ‘I will end up destroying this thing’.” – married PA client, 30-39 years, Nkhotakota

A few PA clients also reported feeling pressure or judgment when being encouraged to take up SI:

> “[Interviewer: What was the nurse saying?] …[she] was saying, ‘Why do you still want us to inject you? Why are you not getting the self-injecting one?… Why are you not injecting yourself?’ … [Interviewer: So what did you say?] I said I have never taken the self-injecting type before.” – married PA client, 20-29 years, Nkhotakota

### Length and feasibility of comprehensive counselling and training

Most providers in this study reported that it takes between 15-20 minutes to counsel and train a woman on DMPA-SC SI. A few others said it could take more than 20 minutes and up to 45 minutes with some clients. Finding this additional time during busy days was not always easy for providers:

> “To explain to the client [about DMPA-SC SI], you need to have uninterrupted time and also sometimes nurses are busy … you need to sit down and start explain[ing] all the procedures up until you are convinced that she can do it. So it’s not that difficult but you just need time to do it” – Facility-based provider, Mzimba South

A few providers mentioned other factors that could influence the length of counselling/training. For example, many providers felt that training a client who had previously used an injectable would be quicker, as she had existing knowledge to draw on, while women with less knowledge of contraception may be more difficult to counsel and train, as they would have more questions to address:

> “Having previous experience [with injectables] does help, for instance, during counselling if we have clients who started the method some time [ago]… you will hear them commenting that they know the stuff … So … this makes the work simple because you know the counselling process will be easy and does not take a lot of time. Whilst if you have new clients it takes long to counsel them” – HSA, Nkhotakota

Providers generally did not mention educational status of the client influencing counselling time, but they did mention age. Most providers felt that adolescents (under 20) required more time to counsel, not necessarily because of difference in understanding but just because they were more likely to be starting from a low baseline of information about contraception:

> “Compared to an older woman, because older women already know the things, and maybe they have already heard the things from somebody else, but mostly the adolescents don’t know most of the information relating to family planning…” – HSA, Zomba

However, a few providers said that it was actually quicker to counsel adolescents because they would pick up new ideas quickly:

> “The understanding of adults is a bit complex… Hence it becomes difficult for them to grasp the information easily as such you have to say it again and again. It’s different from adolescents when you train them, they capture the info easily…” – Facility-based provider, Nkhotakota

A few providers said adolescents’ anxiety about being seen at the facility was a factor in them wanting to avoid group health education sessions or ask for faster counselling:

> “Most youth mostly don’t have time. It happens that sometimes the time we’re giving the talk, they’re rushing to do their own things and they sometimes try to avoid people … they think “If so and so sees me they will report them to their homes.” – Facility-based provider, Zomba

Several providers described their role in counselling on SI becoming quicker and easier over time, as their own experience increased and awareness and acceptance of DMPA-SC at community-level grew:

> “…initially I think for the first three months … we would spend almost 45 minutes… 40 to 45 minutes… but after the first three months we spend 15 minutes” – Facility-based provider, Zomba

Despite the time pressures, most providers took their responsibility for training women in SI very seriously and felt that investing in longer counselling/training facilitated comprehensive understanding, uptake of SI, and reduced likelihood of SI clients encountering problems later. Many providers also clearly articulated that they believed investing that time would increase SI uptake and reduce their workload later, in terms of reducing visits by repeat PA clients.

> “If the provider has counselled the clients so well in details, it becomes so easy for the clients when it comes for them to try to self-inject. But in cases where the provider does counsel the clients in a hurry without checking whether they are clear with the process, [then] it becomes a problem for them when doing the trial injection [practice].” – HSA, Nkhotakota

> “[once trained in SI] … they will just come and collect, and they will not show up again for an entire year. For them [not] to come back, it means [in] the gap they create we can be serving other clients…” - HSA, Zomba

However, a minority of providers admitted that time pressures drove them to take shortcuts in counselling/training, most commonly turning to small-group counselling/training approaches over individual approaches:

> “During busy days, we do shortcuts (laughs)… We wait for at least the women to be 10 or 8 or 7, where you feel that this a good number. You just teach them in one kick [one session] and then give them the methods. But for you to start one-on-one [counselling/training] and [it] being a busy day, it does not work.” – HSA, Nkhotakota

On the clients’ side, SI clients were also more likely than their PA counterparts to say their training had taken a long time, even up to an hour, but typically they were satisfied with this length, as they felt it aided their understanding:

> “[Interviewer: … Did you feel the provider spent enough time training you?] Yes. we spent a lot of time at that place. [Interviewer: you understood what they trained you?] I understood everything they trained me about.” – married SI client, 20-29 years, Nkhotakota

Meanwhile most PA clients who had not taken up SI felt that the counselling and training time they received was too short to fully help them understand:

> “[Interviewer: do you feel you received enough counselling from the provider?] no she was rushing she seemed to have other things to do” – married PA client, 20-29 years, Mzimba South

## Discussion

All providers in this study reported successfully integrating DMPA-SC and the SI option into at least the first stages of their health education and contraceptive counselling approach. During health education sessions, providers tended to focus only on the key benefits of self-injection, to spark client interest in the method, and then follow that up with more in-depth information during individual counselling. In taking this approach, providers seemed to understand that clients’ initial fears at the idea of SI might put them off trying the method and tried to focus on the advantages of the SI option to engage clients in further conversations where they could provide more reassurance than they could in a group health education context.

During role-plays of counselling and SI training, providers generally displayed knowledge and counselling practice on DMPA-SC that aligned with the information from national in-service training curriculum, with a few exceptions where key messages/critical SI steps were missed. Future studies could investigate whether the number of ‘critical steps’ for SI could be reduced without compromising quality of care, and whether this could improve memory retention of the truly critical steps among providers and clients alike. In the meantime, supervision visits for DMPA-SC trained providers should include emphasis on the commonly missed steps/messages outlined in this study to refresh providers’ memories. Use of standardized messaging - such as the intervention by Burke at al. [6] - could be considered to ensure key messages are covered during counselling.

Over time, many providers in this study had developed and innovated their own counselling/training techniques to reassure women sufficiently to take up the SI option. For example, providers relied on 1) comparisons between DMPA-IM and DMPA-SC (and other needle-related concepts) to familiarize the product, 2) showing women the Uniject to reassure them about needle size and simplicity, 3) visualization of the SI steps (either using visual aids or a demonstration with gestures), 4) hearing from other satisfied self-injectors, 5) giving women the opportunity to practice SI (sometimes on demonstration materials but more commonly on themselves under observation) and 6) repeat trainings for the minority of women who needed more time to build confidence.

For their part, clients who successfully took up the SI option typically did so after in-depth counselling and training to build their confidence. SI clients particularly flagged the following techniques as particularly effective at increasing their confidence: the provider taking their time during counselling/training; receiving a visual demonstration; seeing a demonstration; and hearing from other SI clients. Repeated counselling/training was noted to be necessary for a minority of women wanting to try SI, to give them time to get used to the idea and build up their confidence. While providers themselves discussed counselling in-depth on potential side effects, some SI clients in the original sample still wanted more information on these after seeing the ‘best practice’ video. This request from SI clients to cover side-effects comprehensively echoes evidence from other contexts that women really value receiving full and clear information, especially about potential side effects, during counselling. [13]

PA clients who did not take up the SI option tended to report being put off the idea of SI from the start due to fear/doubt in their own ability, and not feeling sufficiently reassured about the messages they may have heard in health education to even learn more about the method. This aligns with evidence from other contexts. [8] For those PA clients who were sufficiently interested to learn more about SI, they were more likely than SI clients to report busy providers unable to take the time to counsel/train them comprehensively, or rushed, incomplete counselling, including lack of visualizations or demonstrations, and lack of testimonials from satisfied self-injectors, before being told to ‘practice’ on themselves. Despite these clear gaps in their counselling experience, in most cases PA clients who did not take up SI tended to blame themselves, particularly mirroring provider language about having ‘failed’. This suggests that in skipping those key steps during counselling and training, providers failed to sufficiently reassure PA clients and even reinforced their lack of confidence.

Providers in this study said that it typically took them around 15-20 minutes (and up to 45 minutes) to train women in SI. This is slightly shorter compared to estimates by providers in previous qualitative studies in Malawi [4], and may reflect providers finding ways to streamline their counselling/training approach over time as they learned the techniques that best reassured women. SI clients in this study tended to report longer counselling (up to an hour in some cases) compared to PA clients, who tended to describe their counselling time as ‘rushed’, suggesting that length and comprehensiveness of counselling may influence confidence to take up SI. Most providers reported investing time to train women in-depth on SI, despite time-pressure and competing priorities, because they felt it was important to ensure clients fully understood the information and because they saw the benefit of reduced workload once women were happily established with using SI. This is similar to previous findings in Malawi. [4] However, some providers in this study reported time pressures leading them to take counselling/training shortcuts; namely, training women in SI in groups, rushing through key messages, and sometimes skipping visualizations or demonstrations. This group training practice was also reported in a recent study with adolescent SI users in Uganda, where around half of the adolescents interviewed would have preferred individual counselling versus around a third reported a preference for group training due to the benefits of peer support and shared experience. [7] Research from the DISC program also found mixed client perspectives on the group training practice in Uganda and Nigeria, with some users emphasizing the need for discretion and confidentiality during training, while other women feeling that peer ‘training’ and other group-based learning about SI may be appealing. [8] Future research should investigate the trade-offs in terms of quality of care between SI training one-on-one or in a group. In addition, peer testimonial approaches should be cognizant of the preference for confidentiality among many SI users, as the researchers from the DISC program note. [8]

Interestingly, most providers and (both PA and SI) clients in this study reported skipping the practice of SI on inanimate objects. While some of this may be due to challenges with DMPA-SC stock and maintaining expensive demonstration materials (i.e. condoms filled with sugar), other providers reported skipping this step because they felt it was not useful to clients. The impact of skipping this step should be explored in future research. However, something that was clear in this study was the negative impact on client confidence of treating women’s first attempt to SI as an ‘exam’. Instead of using narratives implying women ‘passing’ or ‘failing’, providers should be encouraged to help unpack client fears and support clients who still want to self-inject to build confidence over time, as some of their colleagues are already doing. In some cases, multiple sessions may be needed, something that was also found in the DISC program research. [8]

Overall, this study has highlighted the critical role that providers play in ensuring women are confident in SI and has isolated key components of counselling and training that, if skipped, can negatively impact women’s confidence to take up the method.

Key limitations of the study include:

- The sites chosen for the study and the people recruited at these sites may not be representative of the providers/HSAs and clientele at all public sector sites in Malawi. The districts and facilities were sampled randomly to minimize this bias. Results aligning with other similar qualitative studies conducted in Malawi suggest the findings of this study were not unduly affected.
- The perspectives of 15–17-years-old contraceptive users were not included in this study, as the high likelihood of their contraceptive use being covert, combined with the IRB requirement for parental/guardian consent for this population, made their inclusion high-risk for accidental disclosure of their contraceptive use to parents/guardians. It is probable that their perspectives on counselling experience differ from the perspectives of adult clients – as was found in a recent study in Uganda. [7]
- Providers were asked to role-play counselling and SI training sessions with interviewers during the data collection. While these role-plays were likely affected by the knowledge of being observed (Hawthorne effect), they still allowed the interviewers to understand the general approach and content of provider counselling, as well as how it varied between providers.

## Conclusion

Public providers in this study generally demonstrated knowledge and skills on DMPA-SC counselling in line with national in-service training content and shared their examples of techniques for building client confidence with the SI option. However, time- and resource-pressures can lead some providers to deviate from recommended approaches – particularly in terms of grouping women for SI training, and/or skipping visualizations, demonstrations, and the step of practicing on inanimate objects. Clients taking up SI in this study tended to report receiving longer, more comprehensive, and more supportive counselling and training than their PA counterparts who had not taken up SI.

Based on these findings, public providers in Malawi should continue to receive post-training follow up support focused on honing their counselling and training skills, addressing gaps in their knowledge, and sharing best practices for building client confidence to SI, such as visualization, demonstration, and peer testimonials. Use of standardized messaging during counselling could be considered to ensure key messages are covered. Providers should be encouraged to avoid narratives of ‘passing’ or ‘failing’ when training clients, instead focusing on helping to unpack client fears and support clients who still want to SI to build confidence over time.

Future research could explore the impact of removing practice on inanimate objects before proceeding to SI; the impact on quality of care when implementing SI training one-on-one versus in small groups; and explore opportunities to incorporate voluntary SI client testimonials into broader community sensitization activities.

## Data Availability

Anonymized qualitative code reports on themes relating to the topic of the paper have been extracted from Dedoose analysis software and shared as Supporting Information (S2).

## Acknowledgements

The study authors would like to thank the Ministry of Health in Malawi for coordination clearance to work at the study sites; the six data collectors who conducted interviews for this study; and all the providers and clients who contributed their perspectives to this research. We would also like to acknowledge Christina Allain, Irene Obiero, Emma Aldrich and Manish Burman from Clinton Health Access Initiative for their advice and strategic support during the conduct of this study.

**S1. Best practice counselling video script.**

**S2. Malawi DMPA-SC counselling code reports**

## Notes

### Competing Interest Statement

The authors have declared no competing interest.

### Clinical Trial

N/A - not a clinical trial

### Funding Statement

This work was supported by the Bill & Melinda Gates Foundation [Opportunity ID: OPP1195232]. The funders had no role in study design, data collection and analysis, decision to publish, or preparation of the manuscript. Under the grant conditions of the Foundation, a Creative Commons Attribution 4.0 Generic License has already been assigned to the Author Accepted Manuscript version that might arise from this submission.

